# Wastewater genomic surveillance captures early detection of Omicron in Utah

**DOI:** 10.1101/2022.11.24.22282643

**Authors:** Pooja Gupta, Stefan Liao, Maleea Ezekiel, Nicolle Novak, Alessandro Rossi, Nathan LaCross, Kelly Oakeson, Andreas Rohrwasser

## Abstract

Wastewater-based epidemiology has emerged as a powerful public health tool to trace new outbreaks, detect trends in infection and provide an early warning of COVID-19 community spread. Here, we investigated the spread of SARS-CoV-2 infections across Utah by characterizing lineages and mutations detected in wastewater samples. We sequenced over 1,200 samples from 32 sewersheds collected between November 2021 and March 2022. Wastewater sequencing confirmed the presence of Omicron (B.1.1.529) in Utah in samples collected on November 19, 2021, up to ten days before its corresponding detection via clinical sequencing. Analysis of diversity of SARS-CoV-2 lineages revealed Delta as the most frequently detected lineage during November, 2021 (67.71%), but it started declining in December, 2021 with the onset of Omicron (B.1.1529) and its sub-lineage BA.1 (6.79%). Proportion of Omicron increased to ∼58% by January 4th 2022 and completely displaced Delta by February 7th, 2022. Wastewater genomic surveillance revealed the presence of Omicron sub-lineage BA.3, a lineage that is yet to be identified from Utah’s clinical surveillance. Interestingly, several Omicron-defining mutations began to appear in early November, 2021 and increased in prevalence across sewersheds from December to January. Our study suggests that tracking epidemiologically relevant mutations is critical in detecting emerging lineages in the early stages of an outbreak. Wastewater genomic epidemiology provides an unbiased representation of community-wide infection dynamics and is an excellent complementary tool to SARS-CoV-2 clinical surveillance, with the potential of guiding public health action and policy decisions.

## 1. Introduction

Since the beginning of the COVID-19 pandemic, the world has witnessed the evolution of the SARS-CoV-2 virus in real time, with the emergence of new variants and the rapid spread of COVID-19 affecting millions of lives. So far, five variants of concern (VOCs) have been verified by the World Health Organization (WHO): Alpha, Beta, Gamma, Delta and Omicron. Globally, many countries have now lifted COVID-related restrictions, limited vaccine mandates and transitioned to normality. Thus, there remains an increasing concern over the emergence of novel SARS-CoV-2 variants with increased virulence, transmissibility and immune escape potential affecting the efficacy of currently available vaccines (1,2). Over the course of the pandemic, COVID-19 surveillance strategies have relied on testing of symptomatic individuals and their contacts to slow the spread of the pandemic. However, symptoms seem to be less severe in the ongoing Omicron wave, with many affected individuals remaining asymptomatic and often not seeking clinical testing, thereby rendering traditional surveillance systems less reliable. Furthermore, increased adoption of at-home testing results in systematic underreporting and less efficient surveillance. Thus, as the pandemic shifts, relying solely on clinical testing may be insufficient for timely detection of outbreaks and emerging variants. To circumvent these challenges, there is an urgent need to incorporate alternative testing strategies to detect low, circulating levels of SARS-CoV-2 variants.

Wastewater surveillance of SARS-CoV-2 is one such approach that can be used to track transmission and spread of new and existing variants across communities (3–7). Although, wastewater-based epidemiology (WBE) has traditionally been used for surveillance of a variety of targets, including poliovirus (8), antimicrobial resistance genes (9) and monitoring illicit drug-use (10), WBE has recently gained traction as a promising epidemiological tool to track SARS-CoV-2 infections. The Centers for Disease Control and Prevention (CDC) developed a national database - the National Wastewater Surveillance System (NWSS) - in late 2020 to monitor the spread of COVID-19 in the United States (CDC, 2020). WBE of SARS-CoV-2 complements existing clinical surveillance as it can capture both symptomatic and asymptomatic cases. As SARS-CoV-2 is known to be shed in human feces post-infection, usually well before symptoms appear, the viral loads in sewersheds provide a good indicator of SARS-CoV-2 case burden and transmission. Previous studies have found strong correlation between SARS-CoV-2 concentrations in wastewater and reported COVID-19 cases (11–14).

WBE of SARS-CoV-2 is useful in monitoring infections when clinical testing is limited, expensive or overwhelmed due to case surges, especially during the onset of a new variant. It is also not subject to biases related to health care seeking behavior and willingness to be tested. Previous work has shown that WBE could be essential to conducting long-term, population-wide SARS-CoV-2 surveillance in a rapid and cost-effective manner (15). Importantly, WBE can provide an early warning of rising COVID-19 infections in communities (16,17) and could be an important tool for guiding public health interventions in combating the ongoing COVID-19 pandemic.

Despite the advantages of WBE for SARS-CoV-2 surveillance, fewer studies implement genomic analysis of wastewater samples. A wastewater sample represents pooled genomic content from multiple individuals and presents a mix of different viral lineages. Additionally, wastewater samples are often degraded and may contain fragmented viral genomes (18). Thus, it is often challenging to confidently assign lineages in a wastewater sample and assess viral genetic diversity. Consequently, early research efforts have focused mainly on measuring virus titers, usually via qPCR, to track trends in COVID-19 infection (3,12,13) or digital droplet PCR (ddPCR) to characterize variants of interest (VOI) and VOCs (19,20). However, PCR-based approaches are limited in their application due to mutations being shared across VOI/VOCs and ongoing evolution of SARS-CoV-2 virus leading to the occurrence of novel mutations. It also requires continual updating and validation of existing methods to avoid primer dropouts. As ddPCR targets signature mutations, it may miss amplification of other possible variants which may be epidemiologically relevant. To circumvent these issues, recent studies have adapted the use of next generation sequencing approaches to comprehensively scan the entire SARS-CoV-2 genome for potential mutations (21–24). This enables detection of unique and functionally important mutations and VOC-defining clusters of mutations. Novel computational approaches are also being developed to parse the different viral lineages in wastewater samples (21,25,26). Genomic sequencing of SARS-CoV-2 in wastewater offers a universal and powerful tool to assess the prevalence of circulating SARS-CoV-2 variants and help detect new emerging variants in a population.

In this study, we sequenced wastewater samples collected from 32 sewersheds across Utah. Our goal was to track SARS-CoV-2 spread by characterizing lineages and mutations detected in wastewater communities. We also examined whether lineages detected in wastewater were comparable to lineage diversity observed in clinical samples. We collaborated with the Centre for Genomic Pathogen Surveillance to develop an interactive Micoreact dashboard (27) to visualize our wastewater sequencing data and inform local public health investigations. This internal dashboard provides an intuitive summary of wastewater sequencing results to our state epidemiologists and helps in keeping track of various SARS-CoV-2 mutations and associated lineages over time and across various sampling sites.

## 2. Materials and methods

### 2.1. Sample collection

Sewage samples were collected twice per week at 32 wastewater facilities between November 2021 and March 2022 (n = 1,235; Fig. 1; Supplementary Table S1). All samples were raw influent wastewater collected from municipal treatment systems. Estimated resident populations varied widely, from a low of 1,322 to a high of 515,494. While the specific type of sample collected was dependent on sampling equipment available at each partner facility, all but three facilities provided nominally 24-hour time- or flow-weighted composite samples (mean duration of 23.9 hours, range of 16 - 28.3 hours). The remaining three facilities provided either grab samples (Roosevelt City SD) or 6-hour manual composite samples (Snyderville Basin East Canyon and Snyderville Basin Silver Creek; four grab samples collected every two hours over a total of six hours then mixed). Samples were stored in a refrigerator or on ice and transported to the Utah Public Health Laboratory (UPHL) within 24 hours.

**Fig. 1.**
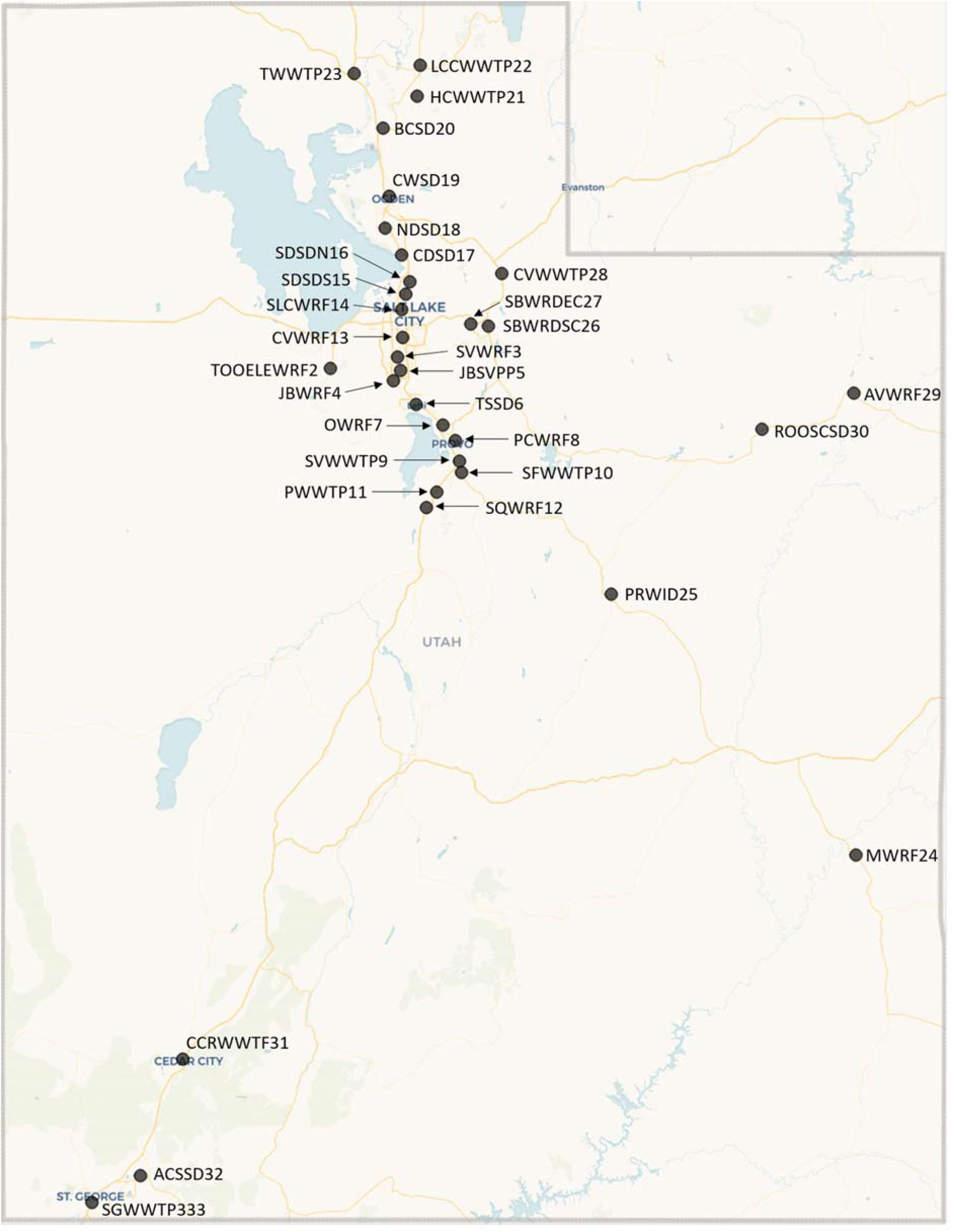
Geographic map showing the 32 sampled wastewater treatment sites in Utah.

### 2.2. Sample processing and qPCR testing

20mL of raw influent wastewater sample was processed following the steps outlined in the Promega Maxwell® RSC Enviro Total Nucleic Acid Kit (Promega Corp.). Briefly, the samples were lysed, the solids were separated, and the samples were buffered. Total nucleic acids were captured on the Promega PureYield™ Midicolumn, purified and eluted to 600 μL. The 600 μL concentrate was then extracted by Promega Maxwell RSC 48 automated extraction and stored at −80°C until RT-qPCR analysis. We conducted RT-qPCR using TaqPath mastermix and the primers and probes contained within the 2019-nCoV CDC EUA kit (28) to detect the presence of the targeted gene sequences (N1 and N2) on a ThermoScientific ABI 7500 Fast instrument. We quantified SARS-CoV-2 RNA against a 5-point standard curve made using serial dilutions of the CDC EUA 2019-nCoV positive control with N1 and N2 analyzed in combination.

### 2.3. Whole-genome Sequencing

We sequenced the SARS-CoV-2 genome using amplicon-based sequencing following the Illumina COVIDSeq™ RUO protocol (Illumina, Inc. San Diego, CA, USA). Library preparation was automated via Tecan liquid handlers and pooled, amplified libraries were loaded onto the S4-flow cell following NovaSeq workflow as per manufacturer’s instructions (Illumina Inc). Dual indexed single-end sequencing with a 75bp read length was performed on the NovaSeq 6000 platform.

### 2.4. Bioinformatics Analysis

After sequencing, we generated FASTQ files from the raw data using the DRAGEN COVIDSeq Test Pipeline (Illumina Inc.) on the Illumina DRAGEN Bio-IT platform. We used viralrecon (v.2.4.1) to analyze the sequencing data and detect SARS-CoV-2 variants (29). Briefly, we performed data QC, trim and quality filtering on raw FASTQ files using fastp. We discarded reads with an average Phred quality score below 30 and length shorter than 50 bp. We mapped the trimmed, human-filtered reads to the SARS-CoV-2 (Wuhan-Hu-1 reference, RefSeq ID: NC_045512.2) genome using bowtie (v.2.4.4) (30). We soft-clipped the ARTIC primers using iVar (v.1.3.1) (31) using default settings. Subsequently, we performed variant calling using iVar and generated consensus sequences using BCFtools (v.1.14) (32). We retained variants with a minimum base quality of 20, allele frequency of 0.25 and coverage of 10 reads. We annotated variants with SnpEff (v.5.0e) (33) and SnpSift (v.4.3) (34). All filtered variants were then compared against a manually curated list of known and epidemiologically important SARS-CoV-2 variants (including VOCs and VOIs) built using the ‘outbreakinfo’ R package (35).

We used Pangolin COVID-19 Lineage Assignment tool (v.4.0) to detect potential SARS-CoV-2 lineages from consensus sequences (36). We obtained a summary of raw reads, sample QC, alignment and variant calling metrics via MultiQC (v.1.11) (37). Unlike a clinical sample, each wastewater sample can contain multiple different SARS-CoV-2 virus lineages shed by many infected individuals. We used Freyja (v1.3.8) (25) to delineate and ascertain the relative proportion of different SARS-CoV-2 lineages in each wastewater sample.

## 3. Results

### 3.1. Viral RNA concentrations

Viral RNA concentrations varied widely over the study period from non-detect to a maximum of over 22,000 copies/mL. The maximum concentrations collected from partner facilities also varied by over an order of magnitude. The highest concentrations were observed at the height of the Omicron wave (mid to late January 2022), while the lowest concentrations were observed shortly thereafter in mid to late March 2022 (Supplementary Fig. S2).

### 3.2. SARS-CoV-2 lineage diversity in wastewater *vs* clinical samples

Of the 1,235 samples collected from 32 sewersheds between November 2021 and March 2022, 1,067 had sufficient RNA for sequencing. The DRAGEN COVIDSeq pipeline successfully generated FASTQ files for 1,034 of those samples. We obtained a total of 9.86 ± 6.08 million raw reads and retained an average of 8.75 ± 5.37 million reads after quality control and read trimming. We found that on average 62.57 ± 25.45% of total reads mapped to the SARS-CoV-2 reference genome, and covered an average 87.37 ± 18.94% of the SARS-CoV-2 genome at 1X across all wastewater samples (Supplementary Fig. S3 and Fig. S4).

We detected a total of 70 different lineages (including sub-lineages and with frequency > 5) from 1,034 consensus sequences following PANGO nomenclature. Analysis of diversity of SARS-CoV-2 lineages revealed Delta as the most frequently detected lineage in our dataset (frequency, n= 1,621, 67.71%) during November, prior to the emergence of Omicron (Fig. 3a). However, we first detected Omicron (B.1.1.529) in a wastewater sample collected on November 19th, 2021 from Hyrum City WWTP (HCWWTP21) at an abundance of 0.1% (Fig. 3b). The first clinical case of Omicron in Utah was reported on November 24th, 2021 with a specimen collection date of November 18th, 2021, though it was travel-related. Two additional clinical cases of Omicron were reported on November 29th, 2021 but community transmission of Omicron in Utah was observed around mid-December (Fig. 3c-d).

We observed a steady decrease in the proportion of the Delta (B.1.617.2) lineages and the onset of Omicron (B.1.1529) and its sub-lineage BA.1 (n=112, 6.79%) in wastewater by December. A simple timeline of the detection of Omicron lineages is shown in Fig. 2. We found BA.1.9 (lineage abundance: 8.35%) from Snyderville Basin Silver Creek (SBWRDSC26) on December 7th, 2021, followed by detection of BA.1.17 (lineage abundance: 4.60%) and BA.1.1.11 (lineage abundance: 27.93%) from Central Valley WRF (CVWRF13) on December 10th (Fig. 4). By December 14th, Omicron spread to two other communities, Snyderville Basin East Canyon (SBWRDEC27; lineages BA.1.1: 15.8%, BA.1.9: 9.11%) and Brigham City (BCSD20; lineage BA.1.9: 9.82%), along with increased prevalence in Snyderville Basin Silver Creek (SBWRDSC26; lineages B.1.1.529: 9.41%, BA.1.9: 52.75% and BA.1.1.16: 14.58%). Over the next two weeks, several BA.1 sub-lineages of Omicron had spread to nearly all communities (28/32), including the then common BA.1.15 (Fig. 4). By January 4th, prevalence of Omicron in wastewater samples had reached 57.58%, and it completely displaced Delta by February 7th, 2022 (Fig. 3a). Between January and March, Omicron accounted for 95% of total lineages in circulation (Fig. 3b), with BA.1 being the most common (n=3,037, 82.93%), followed by BA.2 (n=156, 4.26%) and BA.3 (n=7, 0.19%). Overall, we detected a comparatively rapid shift from the earlier predominant Delta lineage to Omicron in our wastewater data.

**Fig. 2.**
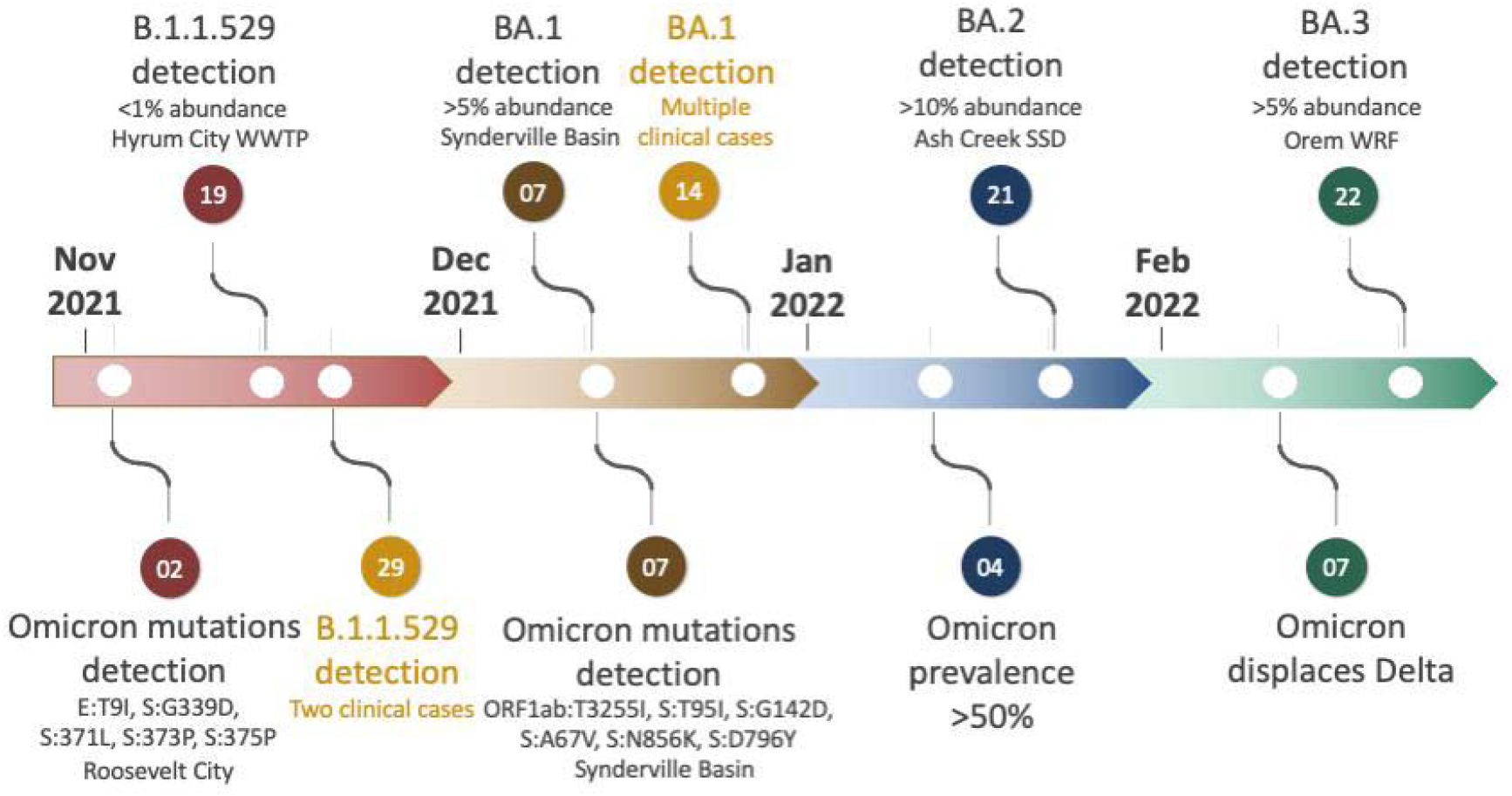
Timeline of the important events in the detection of lineages and key mutations of Omicron from wastewater sequencing.

**Fig. 3.**
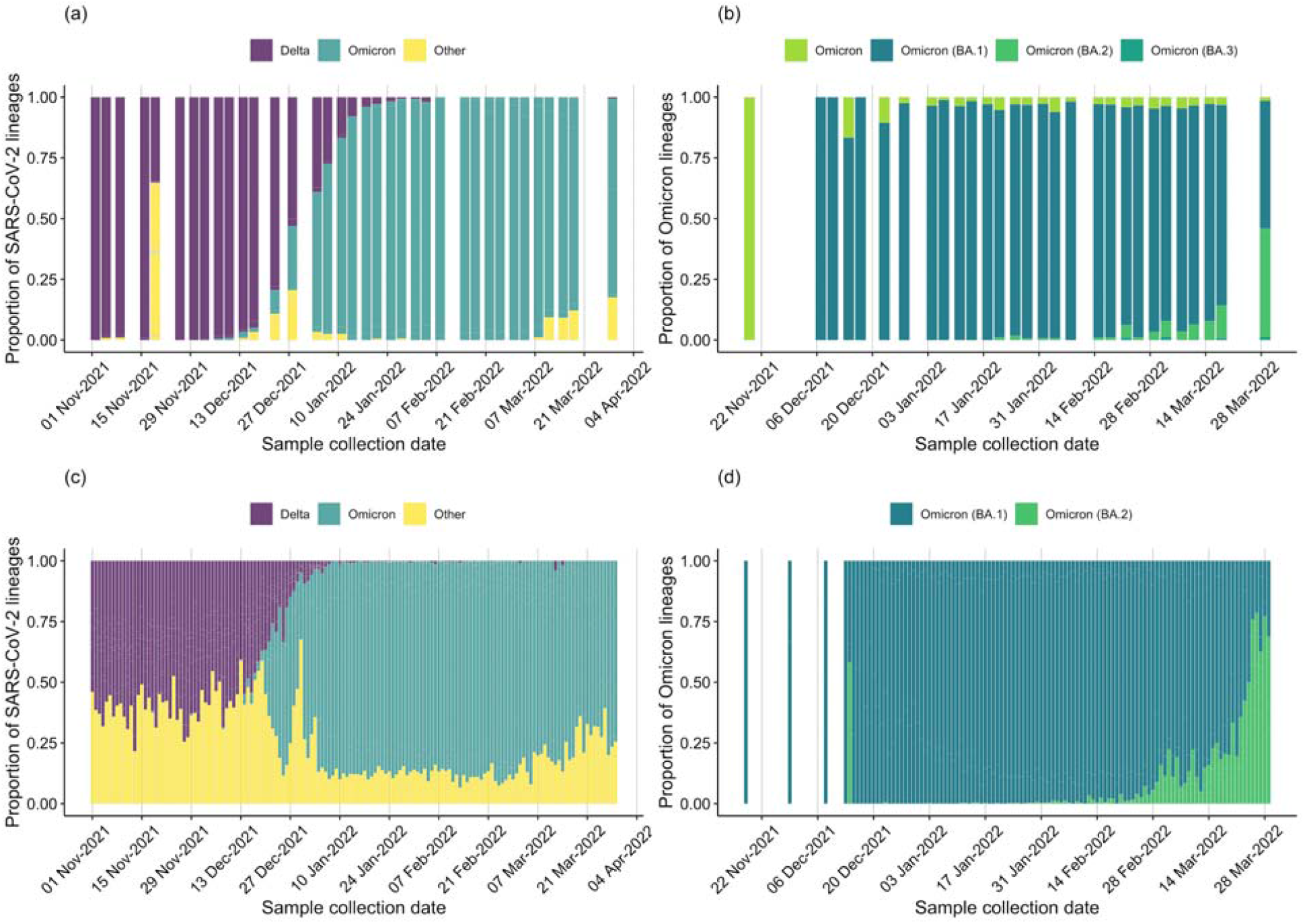
Proportion of all Sars-CoV-2 lineages in a) wastewater samples, c) clinical samples and proportion of Omicron lineages in b) wastewater samples, d) clinical samples from November 2021 to March 2022.

**Fig. 4.**
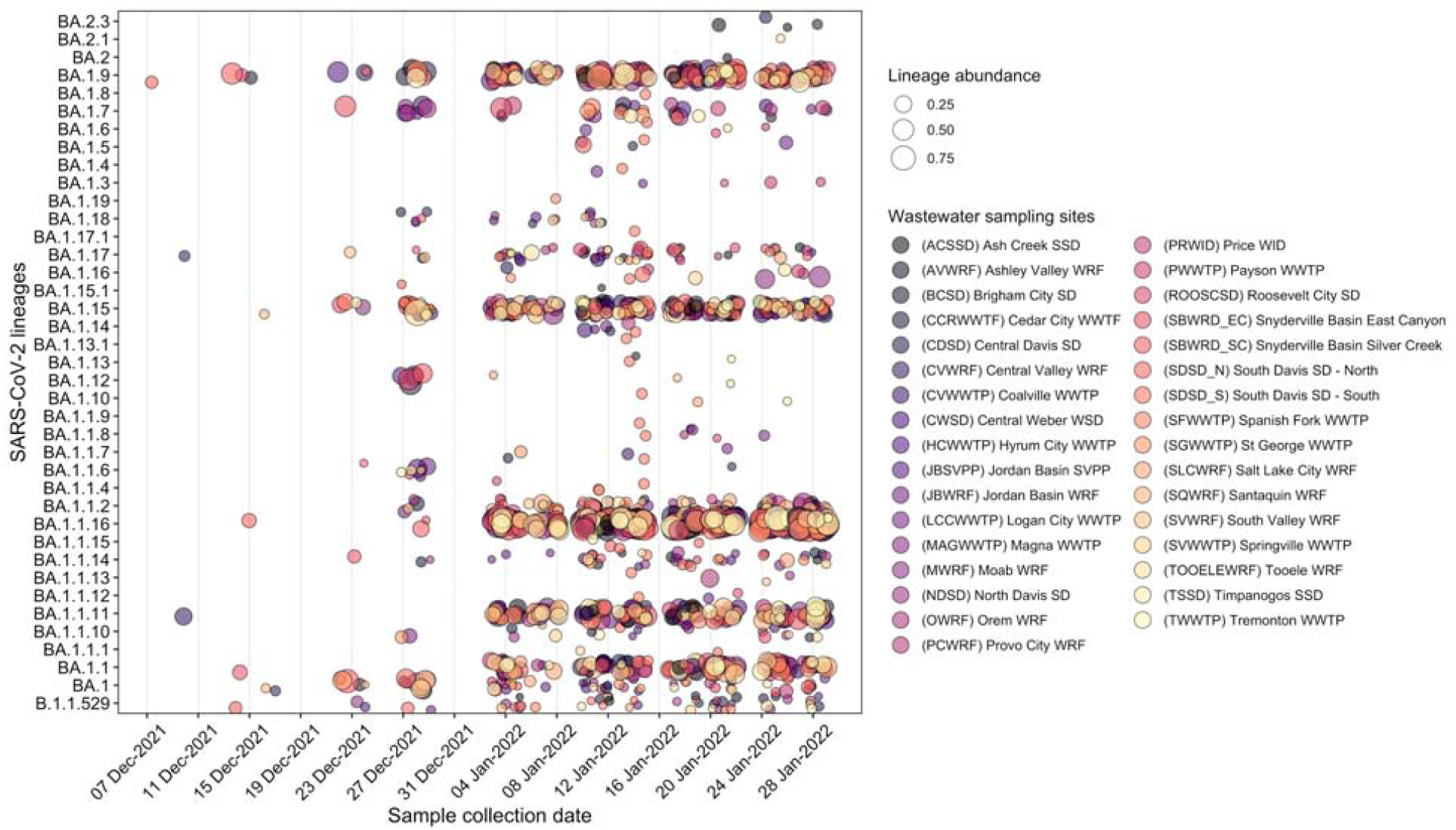
Abundance of Omicron lineages and its sub-lineages in each sewershed between December 2021 and January 2022 to highlight the introduction and spread of Omicron in Utah. Circles are color coded by sewersheds and the size of each circle corresponds to the relative abundance of lineages.

Our wastewater genomic surveillance also identified changes in abundance of other Omicron sub-lineages over time. We first detected the Omicron sub-lineage BA.2 in our wastewater sequencing data from Ash Creek SSD (ACSSD32) on January 21st, 2022 (lineage abundances: BA.2.3, 11.28% and BA.2, 1.79%; Fig. 4). By March 29th, the proportion of BA.2 reached about 45% including its sub-lineages BA.2.1, BA.2.2, BA.2.3, BA.2.4, BA.2.5, BA.2.6, BA.2.7 and BA.2.8. On February 22nd, 2022, we first detected Omicron sub-lineage BA.3 (lineage abundance: 6.69%) in Orem WRF (OWRF7). Interestingly, Omicron BA.3 persisted at low abundance in wastewater samples through March, but the BA.3 sub-lineage has not yet been identified in clinical surveillance. The patterns observed in wastewater sequencing data were reflected in clinical sequencing (Fig. 3c) and resulted in a huge surge in Omicron cases in December-February (Supplementary Fig. S5b). The increased transmission and spike in clinical cases due to Omicron was also evident in wastewater RNA viral loads (Supplementary Fig. S5a) and later also observed in our analysis of the proportion of mutations associated with the Omicron lineage.

### 3.3. Mutational spectra of SARS-CoV-2 in wastewater

We found S:D614G, ORF1ab:T3255I, S:T478K, S:N969K and E:T9I as the most common mutations among all wastewater samples (Supplementary Fig. S6). All five mutations were associated with known VOCs. Notably, all wastewater samples prior to the onset of Omicron (during November 2021) had signature mutations linked to the predominant Delta variant (e.g., S:P681R, S:D950N, T19R), followed by an increase in frequency of mutations shared between Delta and Omicron lineages (e.g., S:L452R, S:T478K; Supplementary Fig. S7).

An in-depth analysis of mutations from wastewater samples revealed an increase in frequency of several Omicron specific mutations (e.g., ORF1ab:T3255I, S:T95I, S:A67V, S:N856K, S:G142D; allele frequency >0.25) in late December 2021 to January 2022 (Fig. 5). Genomic sequencing for wastewater samples collected from Roosevelt City (ROOSCD30) on November 2nd, 2021 confirmed the presence of five signature Omicron mutations (E:T9I, S:G339D, S:371L, S:373P, S:375P). We also identified two Spike protein mutations, A67V and T95I (allele frequency 1.00), specific to the Omicron lineage BA.1 from Provo City (PCWRF8) on November 30th, 2021. Later, we identified six signature mutations of Omicron (ORF1ab:T3255I, S:T95I, S:G142D, S:A67V, S:N856K, S:D796Y) in Snyderville Basin Silver Creek (SBWRDSC26) sampled on December 7th, 2021, supporting the assignment of Omicron BA.1 in our lineage analysis (Fig. 4). We also identified 25 signature mutations of Omicron from Central Valley WRF (CVWRF13) sampled on December 10th, 2021, aligning with its lineage assignment as Omicron BA.1 (Fig. 5).

**Fig. 5.**
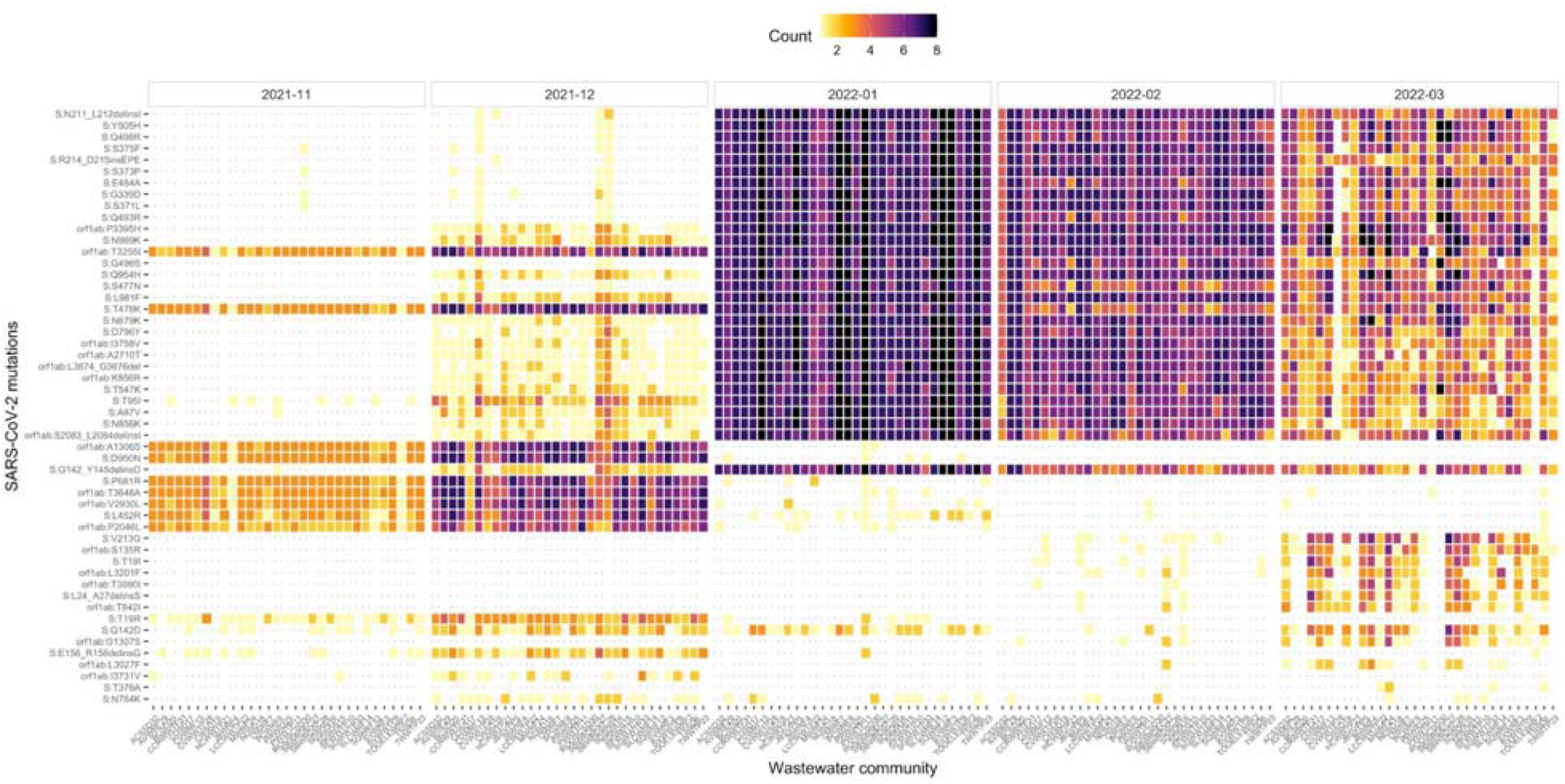
Heatmap showing the temporal changes in the frequency of SARS-CoV-2 amino acid changes in the Spike and ORF1ab genes, detected in each sewershed from November 2021 to March 2022, encompassing the onset of Omicron. Amino acid changes that were detected only once during the study period are not shown. Colors in the heatmap range from cool yellows/reds representing low frequency to warm blues/violets representing high frequency.

## 4. Discussion

As the SARS-CoV-2 virus continues to spread and evolve rapidly to form new variants with increased virulence or immune escape potential, it is important to monitor and characterize VOI/VOCs. This is especially important now when pandemic fatigue has become widespread, clinical testing is reduced and more at-home tests are utilized. These factors all contribute towards delays in detecting new emerging lineages when VOCs may be already circulating in communities. In such scenarios, wastewater genomic surveillance could be essential to capture community transmission of COVID in a rapid, cost-effective and unbiased manner. It can provide a more comprehensive picture of SARS-CoV-2 genomic diversity at the community level than individual level data obtained by clinical sequencing. This will allow timely detection of newly emerging lineages and support public health efforts in mitigating COVID-19 spread (38).

Our wastewater sequencing data provides a first detailed view of the diversity of SARS-CoV-2 lineages circulating between November 2021 and March 2022 in Utah, encompassing the displacement of Delta by Omicron lineages. The trends in SARS-CoV-2 RNA levels across sewersheds accurately captured the surge in clinical cases among Utah’s population (Supplementary Fig. S5). Notably, we found that the prevalence and diversity of SARS-CoV-2 lineages derived from wastewater samples closely aligned with the lineage proportions derived from clinical genomic surveillance. Sequencing of both wastewater samples and clinical samples between December and January revealed a notable increase in the prevalence of Omicron BA.1 lineages and corresponding decrease in the prevalence of Delta lineages (Fig. 3). Omicron BA.1 was the predominant lineage from January through early March, which aligned with its global surge in cases.

One of the main advantages of wastewater surveillance is that it can be used for early detection of emerging lineages and help implement public health interventions to limit variant spread (16,17). In our study, comparing the collection date of Omicron-positive samples from wastewater with the collection dates of samples from clinical genomic surveillance, we detected Omicron lineages in wastewater genomic surveillance on November 19th, 2021, up to 10 days before corresponding detections through genomic clinical surveillance. Notably, the first clinical case was detected in Utah on November 18th, 2021 but was a result of someone with international travel history to a region with high community transmission of Omicron. Two additional clinical cases were identified on November 29th, 2021. Subsequent increases in abundance of Omicron lineages in wastewater on December 7th, 2021 were followed by an increase in the number of clinical cases by December 14th, supporting the week ahead early warning signal detected in wastewater data. The lead time between the detection of a new lineage in wastewater compared to clinical surveillance is still difficult to estimate in advance and has been shown to vary from 5-14 days (12,16,17,39). The lead time can potentially be influenced by environmental factors– temperature, seasons (40), biological factors such as the shedding rates associated with a lineage (41), severity of infection and whether an individual is symptomatic or asymptomatic (39) when contributing to a particular catchment.

In our study, analysis of wastewater samples by Freyja, which provides relative abundance of multiple lineages in a sample, we first detected Omicron (B.1.1.529) from a Hyrum City (HCWWTP) sample collected on November 19th, 2021. However, based on Pangolin lineage assignment on consensus sequence, we first detected the Omicron lineage (BA.1.1) on December 13th, 2021 in Snyderville Basin Silver Creek. This highlights the uncertainty in lineage assignment based on consensus sequences derived from wastewater samples. Pangolin lineage assignment tools are perhaps better at identifying the predominant viral lineage in a community but unlikely to represent the true lineage diversity as is expected in a wastewater sample, an important consideration for future studies.

Throughout the duration of our study encompassing the peak transmission of Omicron, wastewater sequencing identified three sub-lineages of Omicron: BA.1, BA.2 and BA.3. Omicron sub-lineage BA.1 was detected more frequently during our study compared to BA.2 across all sampled communities. By the end of March, as Omicron spread progressed, we observed an increased prevalence of BA.2 (45% prevalence on March 29th), similar to its prevalence in other states in the US. While BA.2 was still dominant across communities, we first detected BA.3 from Orem WRF on February 22nd, 2022. Surprisingly, BA.3 has not been detected in clinical surveillance in Utah. This could possibly be due to its low prevalence as was observed in our wastewater data (Fig. 3b) and has been observed throughout the US (<0.5% prevalence, last checked Sep 19, 2022 on outbreak.info; (42)), suggesting its cryptic circulation in Utah’s wastewater communities. These findings are in agreement with previous studies that highlight the utility of wastewater surveillance in detection of cryptic lineages – novel SARS-CoV-2 lineages that are rarely observed in humans (4,25).

Interestingly, the shift from Delta to Omicron was also quite clear from SARS-CoV-2 mutations in wastewater. We found predominance of Delta mutations in November-early December, followed by an increase in the number of mutations common between Delta and Omicron lineages and then an increase in Omicron defining mutations (Fig. 4 and Supplementary Fig. S7). Thus, our ongoing wastewater genomic surveillance allowed us to detect Omicron lineage even when based on its signature mutations (e.g., S:A67V, S:N856K, S:N969K. E:T9I). Our study suggests that early detection of a lineage is feasible based on signature mutations, however, we caution against the interpretation of lineages based on individual mutations, especially during the early stages of arrival of a new lineage. This is because frequencies of signature mutations associated with a new lineage are generally low, making it difficult to filter out the noise from the actual signal in the sequencing data.

Additionally, confident assignment of lineages for a wastewater sample requires availability of a comprehensive and well-curated lineage database. Novel lineages that have recently diverged from its ancestral lineage and not yet been curated in the SARS-CoV-2 lineage databases (e.g. Pangolin) will be challenging to identify via wastewater sequencing. However, analysis of key mutations from a clinically important region of the SARS-CoV-2 genome (e.g. Spike gene) that can influence transmissibility, immune or vaccine escape could help us identify a potential new lineage in wastewater. Recent studies have reported the presence of several rare, cryptic lineages in wastewater – SARS-CoV-2 lineages that did not match to any known lineages and had not been reported in humans (4,43). These novel lineages contained several rare mutations that had not been identified in routine clinical surveillance and shared many mutations with the circulating VOCs. Routine monitoring of such mutations of concern from wastewater communities could reveal new SARS-CoV-2 lineages before they become epidemiologically relevant and spread to the wider human population, thus guiding public health decision-making.

## 5. Conclusion

Our study illustrates that genomic sequencing of wastewater samples can be used to detect, monitor and provide insights into the diversity of SARS-CoV-2 variants. Of note is the cost-effectiveness of wastewater sequencing as a pooled community sample can be analyzed to monitor disease spread. Wastewater-based genomic epidemiology can be used to track the prevalence of SARS-CoV-2 variants in near real-time, often earlier than information obtained by clinical sample sequencing. Early detection of emerging lineages can provide the time needed to allocate more testing resources to communities showing early detection of an emerging variant and implement public health interventions to limit variant spread. As we move beyond Omicron and new variants emerge, wastewater-based surveillance will be a valuable tool in the timely identification of novel lineages, complementing clinical sequencing and aiding in public health decision-making. While wastewater surveillance has proven to be an essential tool for SARS-CoV-2 genomic surveillance, it would be worthwhile expanding its application to detect other emerging infections and aid surveillance of future emerging infectious diseases.

## Data Availability

The sequence data generated in the present study has been submitted to NCBI Sequence Read Archive and available under BioProject PRJNA812604.

## Acknowledgments

This work was supported by the Centers for Disease Control and Prevention’s (CDC) Epidemiology and Laboratory Capacity for Prevention and Control of Emerging Infectious Diseases (ELC) Enhanced Detection Cooperative Agreement. We thank our partner wastewater treatment facilities across the State of Utah who collected and sent us the wastewater samples. We also thank the UPHL NGS Sequencing Team for conducting the sequencing of wastewater samples.

