## Supplementary Methods, Tables and Figures for "Wastewater genomic surveillance captures early detection of Omicron in Utah"

Flow-Population Normalization

qPCR results were normalized by both the average wastewater flow during the sampling period and the estimated population of the sewershed as follows:

$$((concentration (copies/mL) * 1,000 * flow (liters/day)) \div population) \div1,000,000$$

This yields units of millions of gene copies per person per day. The final division by 1,000,000 is simply to move the results into a more intuitive and easily interpreted range. While not a universal procedure, such normalization is common among wastewater surveillance programs, including at CDC.

**Supplementary Table S1:**

| **Facility Name** | **Facility ID** | **Longitutde** | **Latitude** | **Sample Type** | **Population** | **Average Sample Duration** | **Min Sample Duration** | **Max Sample Duration** | **Sample_matrix** |
| --- | --- | --- | --- | --- | --- | --- | --- | --- | --- |
| (ACSSD) Ash Creek SSD | ACSSD32 | -113.3190102 | 37.19146247 | time-weighted composite | 24,970 | 23.92 | 23.00 | 24.00 | Raw influent wastewater |
| (AVWRF) Ashley Valley WRF | AVWRF29 | -109.5239795 | 40.43959482 | flow-weighted composite | 29,513 | 24.01 | 23.75 | 24.58 | Raw influent wastewater |
| (BCSD) Brigham City SD | BCSD20 | -112.0248354 | 41.50292145 | time-weighted composite | 21,443 | 24.00 | 24.00 | 24.00 | Raw influent wastewater |
| (CCRWWTF) Cedar City WWTF | CCRWWTF31 | -113.0956195 | 37.6836207 | flow-weighted composite | 32,451 | 24.00 | 24.00 | 24.00 | Raw influent wastewater |
| (CDSD) Central Davis SD | CDSD17 | -111.9305849 | 40.99621549 | time-weighted composite | 64,533 | 22.18 | 22.00 | 26.00 | Raw influent wastewater |
| (CVWRF) Central Valley WRF | CVWRF13 | -111.920527 | 40.66256628 | flow-weighted composite | 515,494 | 24.04 | 23.40 | 24.50 | Raw influent wastewater |
| (CVWWTP) Coalville WWTP | CVWWTP28 | -111.3934702 | 40.91979774 | time-weighted composite | 1,322 | 23.00 | 23.00 | 23.00 | Raw influent wastewater |
| (CWSD) Central Weber WSD | CWSD19 | -111.9969163 | 41.23380374 | flow-weighted composite | 208,850 | 23.83 | 22.97 | 24.72 | Raw influent wastewater |
| (HCWWTP) Hyrum City WWTP | HCWWTP21 | -111.8445487 | 41.63252893 | time-weighted composite | 9,095 | 24.03 | 23.87 | 24.42 | Raw influent wastewater |
| (JBSVPP) Jordan Basin SVPP | JBSVPP5 | -111.9356275 | 40.53223557 | flow-weighted composite | 89,507 | 23.97 | 22.83 | 24.67 | Raw influent wastewater |
| (JBWRF) Jordan Basin WRF | JBWRF4 | -111.9709924 | 40.48935379 | flow-weighted composite | 94,484 | 23.77 | 22.67 | 24.17 | Raw influent wastewater |
| (LCCWWTP) Logan City WWTP | LCCWWTP22 | -111.8291693 | 41.75300043 | flow-weighted composite | 94,005 | 23.38 | 16.00 | 28.32 | Raw influent wastewater |
| (MWRF) Moab WRF | MWRF24 | -109.5098592 | 38.54116922 | flow-weighted composite | 9,896 | 24.00 | 24.00 | 24.00 | Raw influent wastewater |
| (NDSD) North Davis SD | NDSD18 | -112.0192526 | 41.10596782 | flow-weighted composite | 228,700 | 24.00 | 23.98 | 24.02 | Raw influent wastewater |
| (OWRF) Orem WRF | OWRF7 | -111.7083169 | 40.31029221 | flow-weighted composite | 112,901 | 23.99 | 23.52 | 24.57 | Raw influent wastewater |
| (PCWRF) Provo City WRF | PCWRF8 | -111.6457205 | 40.24569662 | flow-weighted composite | 102,624 | 24.08 | 23.37 | 24.73 | Raw influent wastewater |
| (PRWID) Price WID | PRWID25 | -110.813236 | 39.61794056 | time-weighted composite | 17,312 | 23.98 | 22.33 | 25.25 | Raw influent wastewater |
| (PWWTP) Payson WWTP | PWWTP11 | -111.7405997 | 40.03537716 | flow-weighted composite | 21,231 | 24.00 | 24.00 | 24.00 | Raw influent wastewater |
| (ROOSCSD) Roosevelt City SD | ROOSCSD30 | -110.009319 | 40.29239498 | grab | 6,790 | N/A | N/A | N/A | Raw influent wastewater |
| (SBWRD_EC) Snyderville Basin East Canyon | SBWRDEC27 | -111.5586783 | 40.71986302 | manual composite | 23,304 | 6.04 | 6.00 | 6.25 | Raw influent wastewater |
| (SBWRD_SC) Snyderville Basin Silver Creek | SBWRDSC26 | -111.4656292 | 40.71084446 | manual composite | 6,390 | 6.01 | 5.43 | 6.30 | Raw influent wastewater |
| (SDSD_N) South Davis SD - North | SDSDN16 | -111.8859568 | 40.88807396 | flow-weighted composite | 84,981 | 25.13 | 23.25 | 26.37 | Raw influent wastewater |
| (SDSD_S) South Davis SD - South | SDSDS15 | -111.9087842 | 40.83866066 | flow-weighted composite | 21,872 | 24.06 | 23.83 | 24.75 | Raw influent wastewater |
| (SFWWTP) Spanish Fork WWTP | SFWWTP10 | -111.6106251 | 40.11395836 | time-weighted composite | 53,313 | 23.95 | 23.25 | 24.00 | Raw influent wastewater |
| (SGWWTP) St George WWTP | SGWWTP33 | -113.5770535 | 37.07693932 | time-weighted composite | 92,047 | 24.18 | 23.58 | 25.23 | Raw influent wastewater |
| (SLCWRF) Salt Lake City WRF | SLCWRF14 | -111.9312436 | 40.77763247 | flow-weighted composite | 209,645 | 23.17 | 20.78 | 26.15 | Raw influent wastewater |
| (SQWRF) Santaquin WRF | SQWRF12 | -111.7941195 | 39.97082251 | time-weighted composite | 12,510 | 24.00 | 24.00 | 24.00 | Raw influent wastewater |
| (SVWRF) South Valley WRF | SVWRF3 | -111.9507752 | 40.58533129 | time-weighted composite | 254,971 | 23.94 | 23.48 | 24.45 | Raw influent wastewater |
| (SVWWTP) Springville WWTP | SVWWTP9 | -111.6204876 | 40.16382705 | time-weighted composite | 35,840 | 24.00 | 24.00 | 24.00 | Raw influent wastewater |
| (TOOELEWRF) Tooele WRF | TOOELEWRF2 | -112.3082523 | 40.53938919 | flow-weighted composite | 36,244 | 23.99 | 23.00 | 24.67 | Raw influent wastewater |
| (TSSD) Timpanogos SSD | TSSD6 | -111.8497104 | 40.39361735 | time-weighted composite | 253,112 | 24.07 | 23.55 | 24.63 | Raw influent wastewater |
| (TWWTP) Tremonton WWTP | TWWTP23 | -112.1841168 | 41.72197719 | flow-weighted composite | 12,451 | 24.00 | 24.00 | 24.00 | Raw influent wastewater |

**Supplementary Figures:**

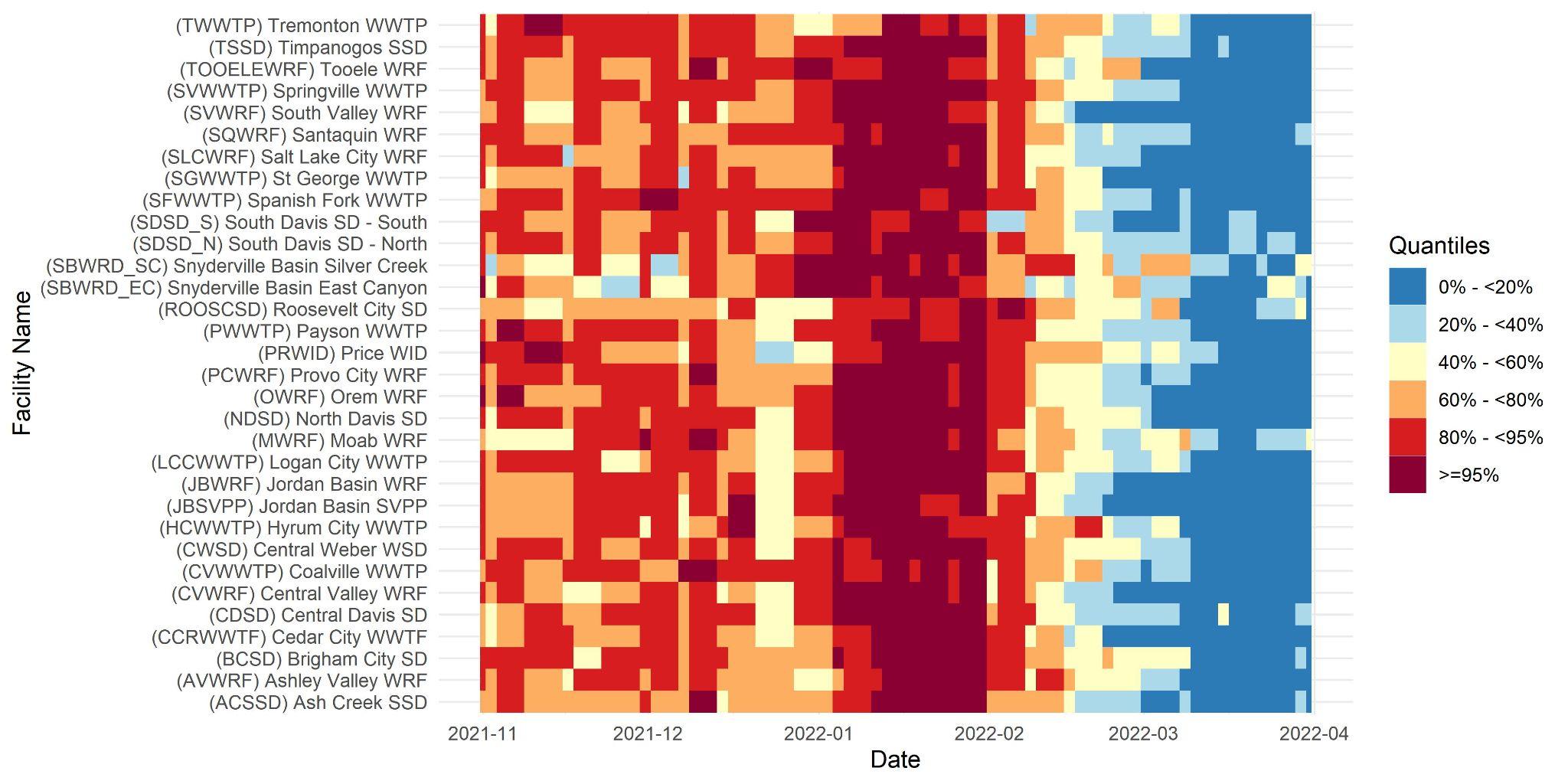

Supplementary Fig. S2. Heatmap of wastewater concentration quantiles. SARS-CoV-2 concentrations in wastewater (normalized by flow and population) were categorized into quantile bins. Bin thresholds are standard quintiles with the addition of a 95%+ bin to better emphasize the highest concentrations. The numeric thresholds were calculated on a site-specific basis on all data from July 1, 2020 to September 22, 2022, although only data relevant to the current manuscript is displayed.

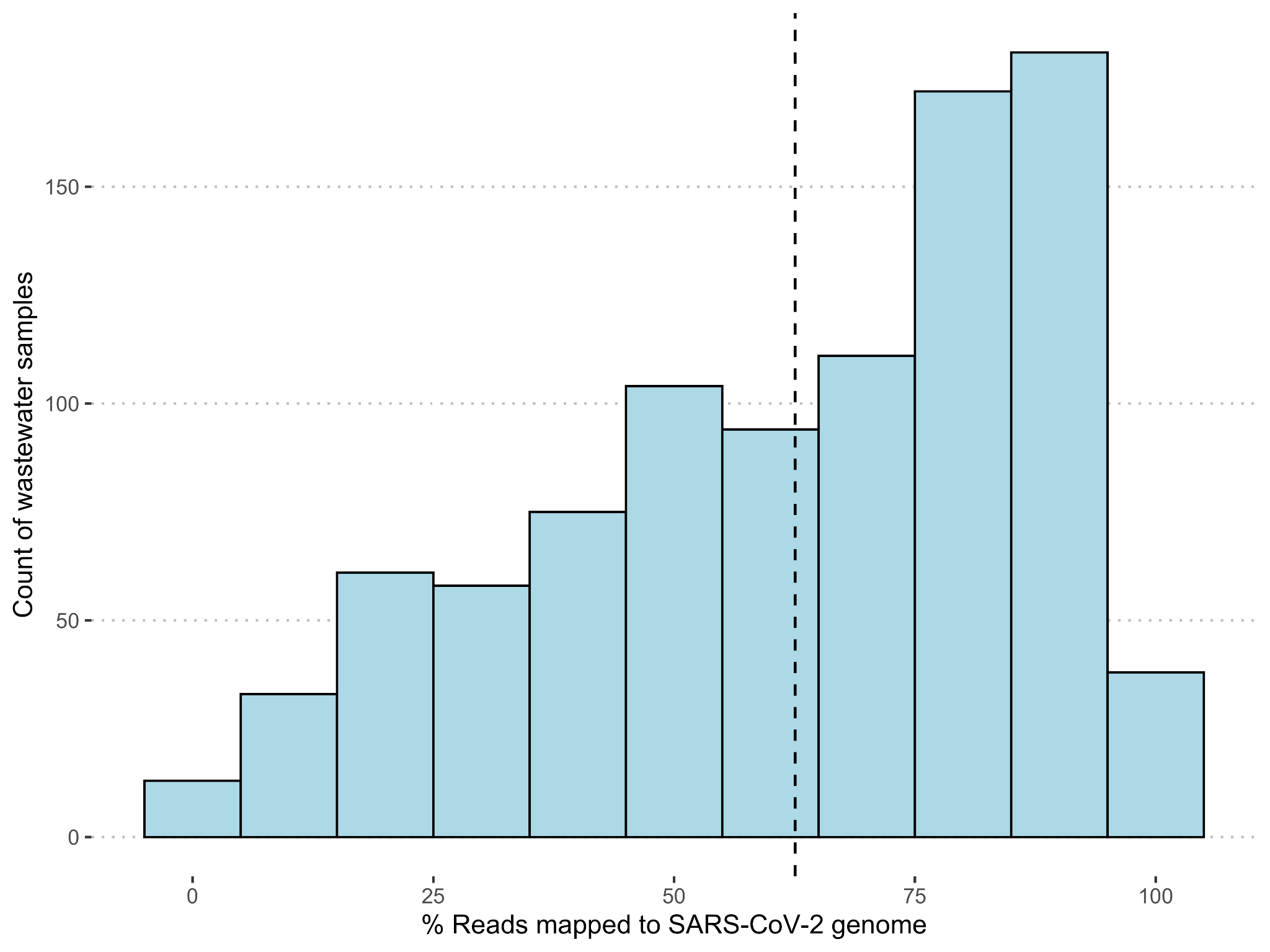

Supplementary Fig. S3. Histogram of the number of wastewater samples and their percent reads mapped to the SARS-CoV-2 genome. The black dashed line indicates the mean percentage of mapped reads.

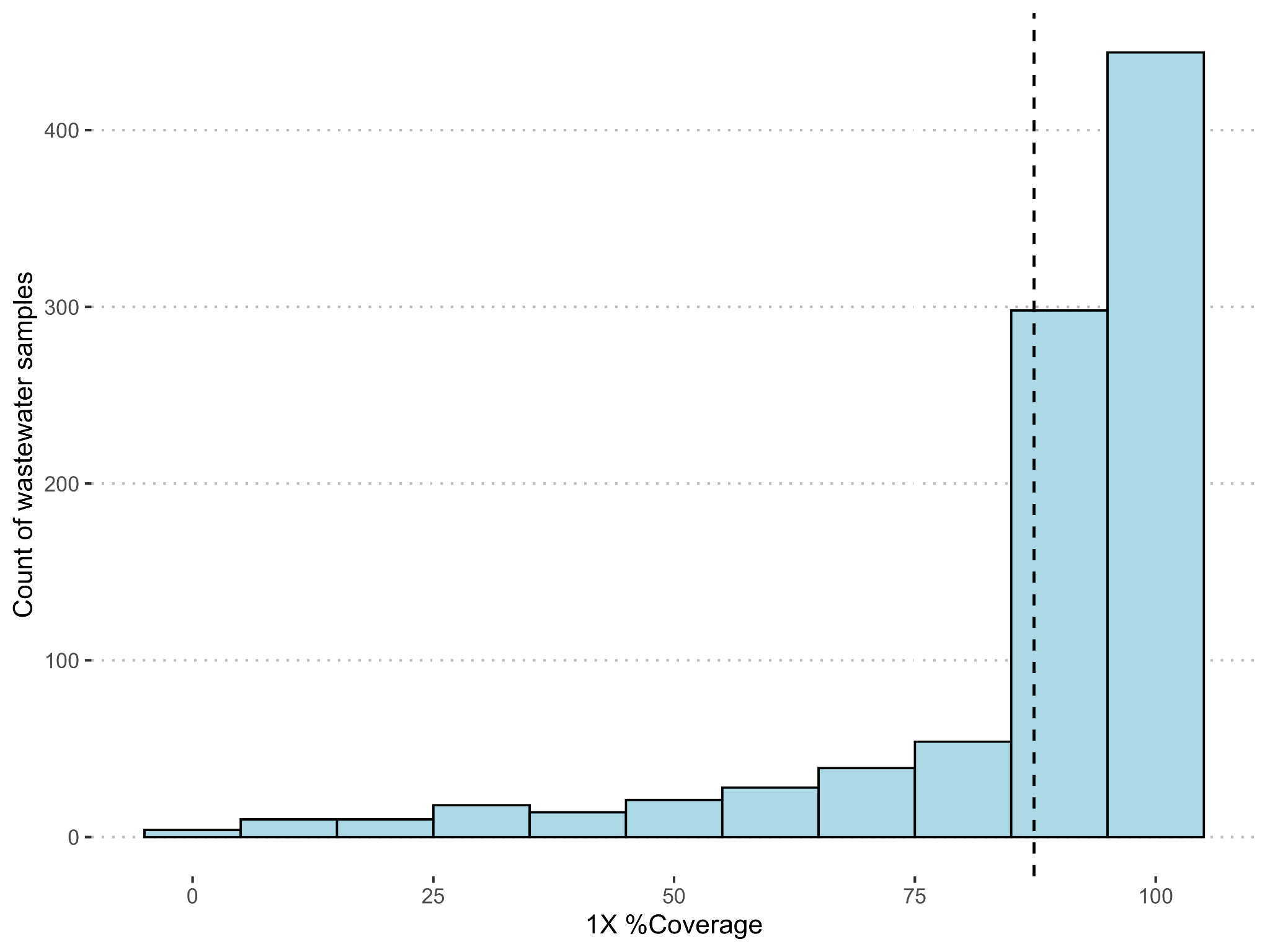

Supplementary Fig. S4. Histogram of the number of wastewater samples and their 1X coverage. The black dashed line indicates the mean 1X coverage.

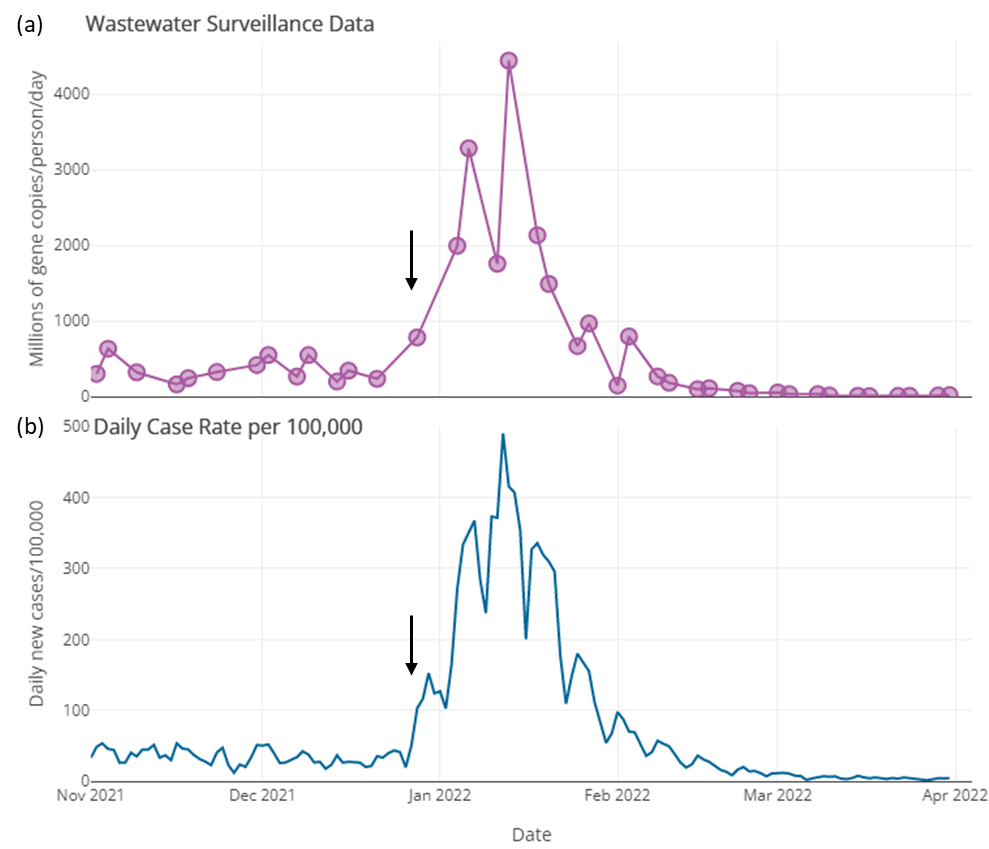

Supplementary Fig. S5. Example wastewater surveillance data and sewershed-associated case rates. The displayed data are from the (CVWRF) Central Valley WRF partner facility, and are generally representative of data from the time period included in the study. Vertical black arrows indicate the approximate start of the Omicron wave in late December 2021.

(a) Wastewater surveillance data. Data generated by UPHL in gene copies/mL of raw wastewater was normalized by estimated population and average flow during the sampling period (details in Supplementary methods). (b) Daily sewershed associated case rates per 100,000. All COVID-19 cases in Utah are geocoded by sewershed.

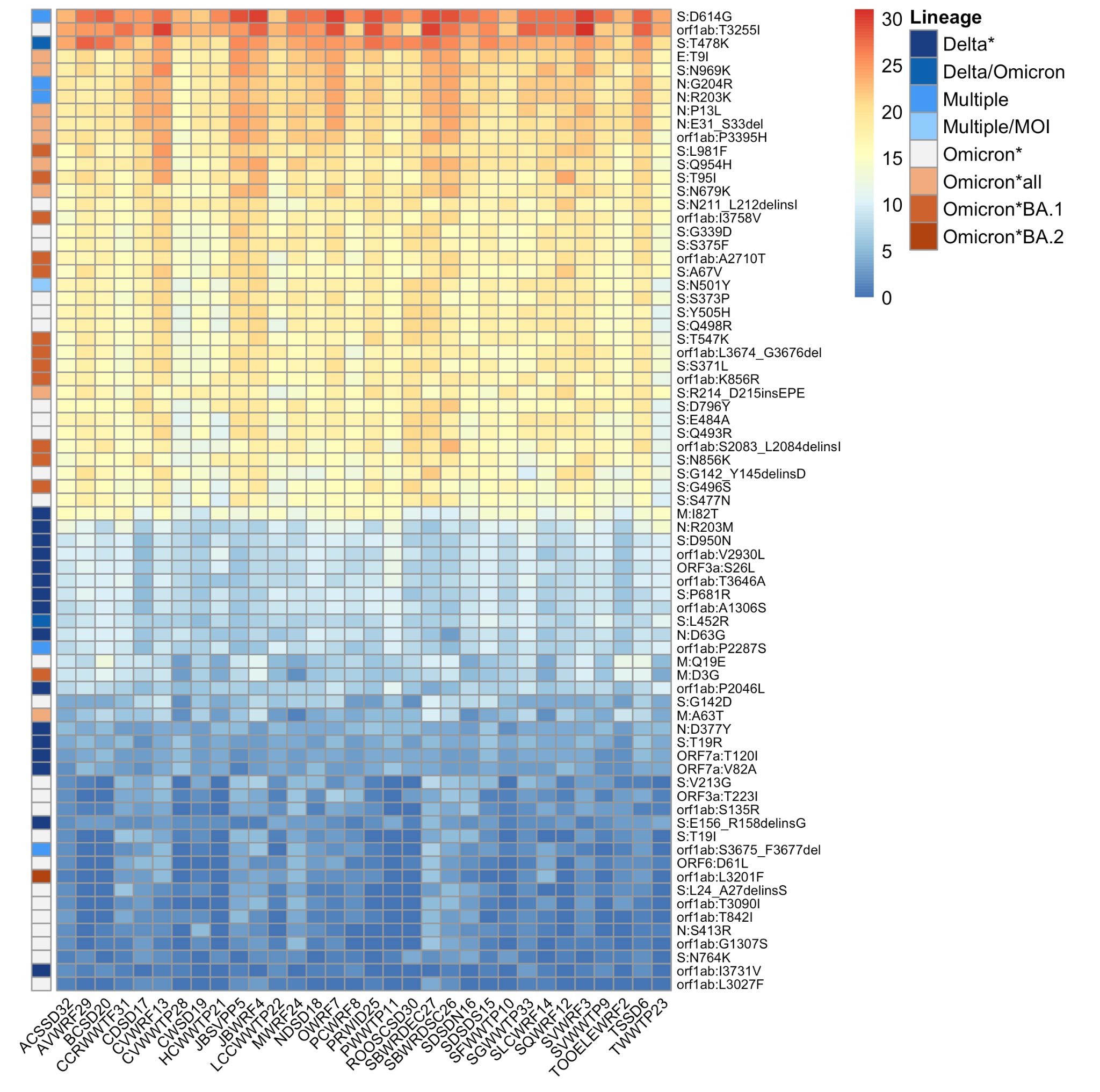

Supplementary Fig. S6. Heatmap of the frequency of all amino acid changes during November 2021 to March 2022 for each sewershed. Colors in the matrix range from cool blues representing low frequency to warm orange representing high frequency. The amino acid changes specific to a SARS-CoV-2 lineage are color coded on the left bar of the heatmap – ‘Multiple’ refers to mutations shared between multiple SARS-CoV-2 lineages and ‘Multiple/MOI’ refers to ‘mutations of interest’ shared between multiple SARS-CoV-2 lineages. Data shown here has been truncated to amino acid changes that were detected 10 or more times during the study period.

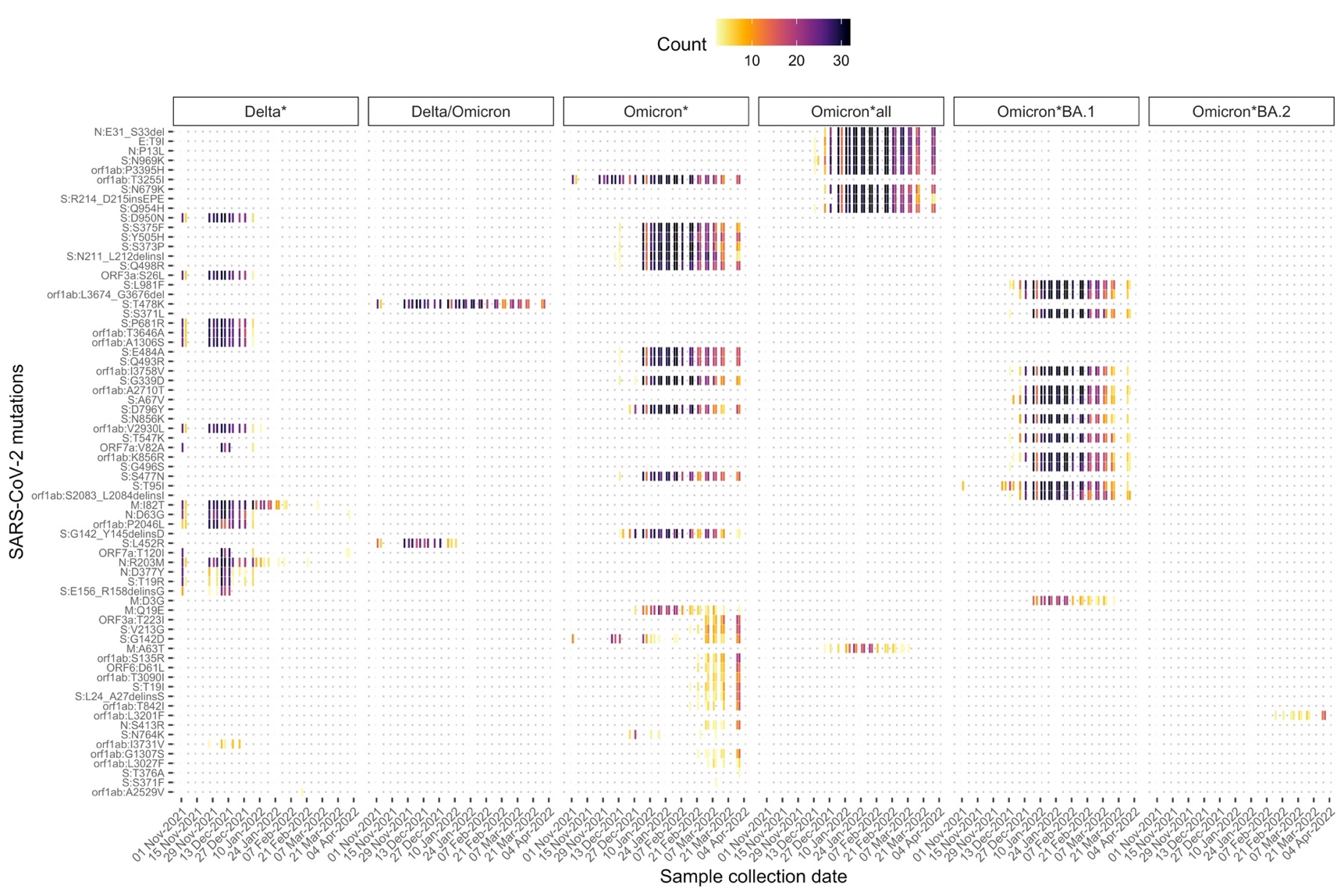

Supplementary Fig. S7. Heatmap showing the temporal changes in the frequency of SARS-CoV-2 amino acid changes from November 2021 to March 2022. Amino acid changes that were detected only once during the study period are not shown. Colors in the matrix range from light yellow/reds representing low frequency to dark blues/violet representing high frequency.
